# Succinylcholine is Equally Efficient as Rocuronium Bromide in terms of Major Adverse Cardiac Events (MACE)During Off Pump Coronary Artery Bypass Surgery: A Single Centre Study

**DOI:** 10.1101/2021.06.21.21259242

**Authors:** Nikhil Mudgalkar, venkata ramana kandi

## Abstract

**Introduction:** Rapid and safe endotracheal intubation is of paramount importance in the general anaesthesia practices. Safety of such practices while performing surgical procedures in people with critical coronary lesions assumes increased significance.The use of succinylcholine and rocuronium are common but the association with the application of these medications and concomitant haemodynamic changes on major adverse cardiac events (MACE) has not been adequately studied. The aim of this study is to assess the safety and efficacy of succinylcholine in comparison with rocuronium for MACE in cardiac surgical population.

**Methods:** Retrospective analysis of data collected from administrative and surgical databases of a tertiary care centre. The patients were divided in two groups,wherein the Group A constituted patients who belonged to succinylcholine and Group B represents the patients who were treated with rocuronium.The baseline demographic characteristics, MACE including intubation difficulty score and Cormack Lahne grade of intubation were recorded.

**Results:** A total of 134 patients were included in the study. Baseline characters were similar in both the groups. There were 2 deaths in the succinylcholine group while 3 in the rocuronium group. The MACE was not statistically significant (p= 0.0505) in both groups. Cormack Lahane scale and intubation difficulty scale were similar in both groups.

**Conclusion:** Succinylcholine was found to be equally efficient and safe in comparison with rocuronium in terms of MACE during coronary artery bypass surgery.

## Introduction

The general anaesthesia practices that include rapid and safe endotracheal intubation is of paramount importance during critical cardiac surgical procedures. Securing the patients airway smoothly and quickly, minimises the chances of regurgitation and aspiration of gastric contents^1,2^.Several studies have been performed on the application of different drugs^3^, varied doses, and timings of administration while performing intubation in different sets of patients. Although cardiac surgery per se is not a risk factor for difficult intubations, there is a higher incidence of difficult intubation ^4,5^ in cardiac surgical population. Factors that may contribute to adverse events include patients with prior coronary artery disease, obesity, diabetes, among others.The duration of intubation is also an important aspect that may contribute to complications during intubation. This may result in the development of cardiac arrest among critical cardiac surgery patients. Rocuronium bromide and succinylcholine are rapidly acting muscle relaxants that act as neuromuscular blockers and are used to manage cardiac surgical patients. Though rocuronium is perceived to be more cardio stable as compared to succinylcholine, rapid clearance of succinylcholine makes it an alternate and a reasonable choice. In this context, and because no studies have been performed to assess the perioperative outcomes of cardiac surgical patients when treated with these two medications, we attempted to study the impact of these two medications on MACE in patients posted for elective coronary artery bypass graft.

## Methods

This retrospective study was undertaken at tertiary care hospital based in rural India during the period between January 2020 and December 2020 and included 146 adult patients who underwent elective or emergency coronary artery bypass graft surgery during the study period.

In all, 146 adult patients of either sex in the age group of 18-80 years were operated for coronary artery bypass graft that required endotracheal intubation. The data was captured from the Institutional administrative surgical and anaesthesia database. The Patients were divided into two groups: Group A (succinylcholine, n=98) and Group B (rocuronium, n=36). The choice of relaxant was made by the primary anaesthesiologist who intubated the patient.

Standard operating protocol of the institute were applied to the patient^6^. The patients in Group A received 1.5mg/kg body wt. of succinylcholine to facilitate endotracheal intubation. The Group B included patients who received 0.9mg/kg body wt. of rocuronium bromide to aid endotracheal intubation.

The primary end point of the study was incidence of MACE in perioperative period (30days) that included episodes of non-fatal cardiac arrest, chaotic rhythm, acute myocardial infarction, congestive heart failure, cardiac arrhythmia, angina, and death^7^.The secondary end points included wererelated to the intubation related issues associated with the use of drugs likeCormack-Lehane grade of glottis visibility obtained by direct laryngoscopy (range, 1-4; 1 indicates full view of glottis; 2, partial view of glottis; 3, only epiglottis seen; and 4, neither glottis nor epiglottis seen), the overall difficulty of the intubation process measured by the intubation difficulty scale score (range, 0 to infinite; score >5 indicates difficult intubation)^8^.

### Statistical Analysis

The Statistical Package for Social Sciences (SPSS) version 16.0.0 for Windows (SPSS Inc., Chicago, IL, USA) and open epi software by CDC version 3.01 updated 2013/04/06 were used to analyse data.

## Results

Thefinal study cohort consisted of 134 patients after excluding 12 patients whose records wereincomplete.The baseline demographic characteristics revealed similar mean age in both succinylcholine(64.02+-12.78) and rocuronium (62.22+_18.16) groups. Both the groups revealed similar number of grafts (2.32+-0.57, 2.34+-0.44, p=0.8491). More than 60% of the study population were males, had diabetes, and hypertension. Moderate ejection fraction was noted in more than 60% of the study subjects. MACE wasobserved to be similar in both the groups as shown in Table 1. The Table 2 reveals the Cormack-Lehane grade of glottis visibility obtained by direct laryngoscopy and intubation difficulty score. The p-values obtained were found statistically insignificant for both the variables.

**Table 1:**
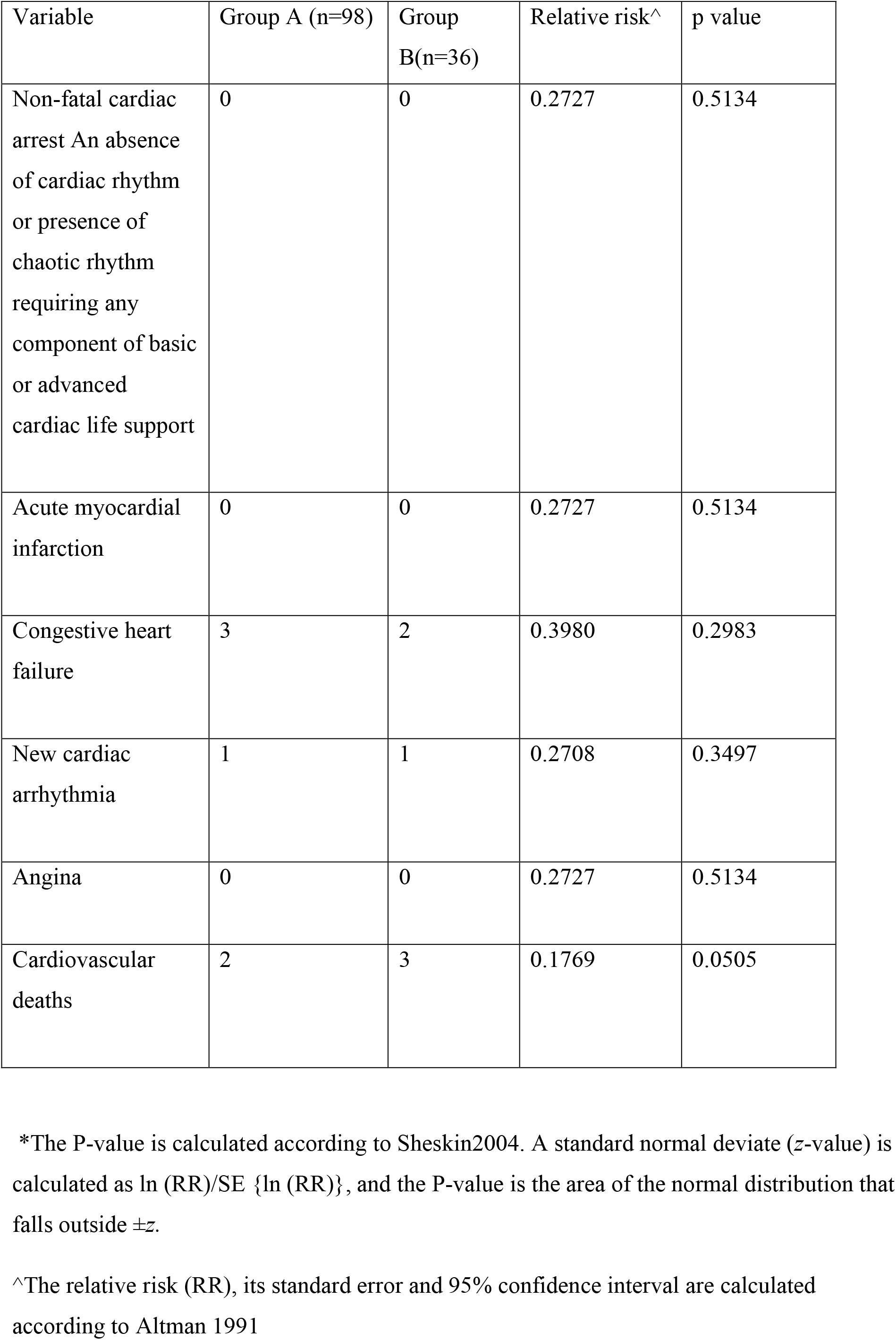
Incidence of MACE in both groups.

| Variable | Group A (n=98) | Group B(n=36) | Relative risk<sup>^</sup> | p value |
| --- | --- | --- | --- | --- |
| Non-fatal cardiac arrest An absence of cardiac rhythm or presence of chaotic rhythm requiring any component of basic or advanced cardiac life support | 0 | 0 | 0.2727 | 0.5134 |
| Acute myocardial infarction | 0 | 0 | 0.2727 | 0.5134 |
| Congestive heart failure | 3 | 2 | 0.3980 | 0.2983 |
| New cardiac arrhythmia | 1 | 1 | 0.2708 | 0.3497 |
| Angina | 0 | 0 | 0.2727 | 0.5134 |
| Cardiovascular deaths | 2 | 3 | 0.1769 | 0.0505 |

\*The P-value is calculated according to Sheskin2004. A standard normal deviate (z-value) is calculated as $\ln(RR)/SE\{\ln(RR)\}$ , and the P-value is the area of the normal distribution that falls outside $\pm z$ .
<sup>^</sup>The relative risk (RR), its standard error and 95% confidence interval are calculated according to Altman 1991

**Table 2:** Intubation difficulty score in both groups:

| Variable | Group A (n=98) | Group B (n=36) | p value |
| --- | --- | --- | --- |
| Cormack-Lehane grade of glottis visibility obtained by direct laryngoscopy | 2.13+_0.8 | 2.11+_0.76 | 0.7980 |
| Intubation Difficulty Scale score | 2.01+-1.1 | 2.07+-0.88 | 0.2022* |

\*P value is calculated from 2 sample independent t test.
P values greater than 0.05 are non-significant.

## Discussion

The documented side effects of succinylcholine are vast with the spectrum ranging from simple postoperative myalgia to life-threatening hyperkalaemia and anaphylaxis. It was previously suggested that succinylcholine would not have been approved in today’s era by the regulatory agencies if it were being investigated as a new drug in the current era^9^. Despite these observations, the use of succinylcholine has not diminished, majorly due to the non-availability of suggamadex in countries like India and due to increased difficult intubation cases presenting for cardiac surgeries^10^.In such scenario, we assessed the MACE associated with the use of succinylcholine in a tertiary care centre that caters the needs of rural population. The incidence of MACE in succinylcholine and rocuronium groups was same indicating non inferiority of succinylcholine for cardiac surgical population compared with rocuronium.

The major concern while using succinylcholine is hyperkalaemia which can lead to fatal cardiac arrest. This can be addressed by regular monitoring of serum potassium levels at pre intubation and thereafter which is routinely performed in cardiac surgical population.

Hemodynamic changes associated with succinylcholine are tachycardia, hypertension and increase in rate pressure product. The present study was done to know whether these changes are associated with mortality in patients^11^. This implicates that even though MACE can happen with succinylcholine, they do not culminate into mortality difference at 30 days and is similar as with Rocuronium.

The patients in cardiac surgery are prone to complications arising from difficult intubation procedures^4^.The incidence of difficult intubation seems 24% in cardiac surgical population compared to 14 % in general population. The nature of critical lesions in coronaries is such that a slightest difficulty or repeated attempts with intubation may lead to fatal cardiac arrest. Suggamadex is still not available in our setups very frequently. Many Cardiac surgical units, especially in the rural areas are managed by a single cardiac surgeon and alone cardiac anaesthetist where it would be difficult for theanaesthetist to face “can’t intubate” problem. So, it is prudent to use succinylcholine more frequently in resource poor centres.

## Conclusion

Succinylcholine is equally efficient and safe when compared to rocuronium for cardiac surgical patients posted for coronary artery bypass surgery in terms of MACE. Therefore, anaesthesiologists may prefer to use succinylcholine more frequently and particularly when difficult intubation scenario arises.

## Limitations

This is a single-centre research. It’s possible that generalising to a wider population won’t be accurate until additional studies with many centres conduct comparable research.The power of study is 80 % with the number of participants enrolled.(Kelsey et al with continuity correction)

## Data Availability

data is available with primary author upon request

## References

1. Xu Z, Ma W, Hester DL, Jiang Y. Anticipated and unanticipated difficult airway management. Curr Opin Anaesthesiol. 2018; 31:96–103. doi: 10.1097/ACO.0000000000000540. PMID: 29176376.

2. Kamalakar K, Vamshidhar B, Saketh K V. Submental Intubation - An alternative airway control. Perspectives in Medical Research 2018; 6:9–12

3. Stollings JL, Diedrich DA, Oyen LJ, Brown DR. Rapid-sequence intubation: a review of the process and considerations when choosing medications. Ann Pharmacother. 2014; 48:62–76. Epub 2013 Nov 4. PMID: 24259635.

4. Borde DP, Futane SS, Daunde V, Zine S, Joshi N, Jaiswal S, et al. Are cardiac surgical patients at increased risk of difficult intubation? Indian J Anaesth. 2017; 61:629–635. PMID: 28890557; PMCID: PMC5579852.

5. Krebs ED, Hawkins RB, Mehaffey JH, Fonner CE, Speir AM, Quader MA, et al. Is routine extubation overnight safe in cardiac surgery patients? J Thorac Cardiovasc Surg. 2019; 157:1533-1542.e2. Epub 2018 Nov 14. PMID: 30578055; PMCID: PMC6431279.

6. Mudgalkar N. Anaesthesia for off pump coronary artery bypass-Recent updates.Perspectives in Medical Research 2017; 5:1-2 DOI: 10.47799/pimr.0502.01

7. Sabaté S, Mases A, Guilera N, Canet J, Castillo J, Orrego C, et al, ANESCARDIOCAT Group. Incidence and predictors of major perioperative adverse cardiac and cerebrovascular events in non-cardiac surgery. Br J Anaesth. 2011; 107:879–90. Epub 2011 Sep 2. PMID: 21890661.

8. Adnet F, Borron SW, Racine SX, Clemessy J, Fournier Plaisance P, et al. The Intubation Difficulty Scale (IDS): Proposal and evaluation of a new score characterizing the complexity of endotracheal intubation. Anesthesiology 1997; 87:1290–7.

9. Blobner M, Hunter JM. Another nail in the coffin of succinylcholine? Br J Anaesth. 2020; 125:423–425. Epub 2020 Jul 15. PMID: 32682561.

10. Kopman AF, Brull SJ. Not another requiem for succinylcholine. Comment on Br J Anaesth 2020; 125:423-5. Br J Anaesth. 2020;125: e349–e350. Epub 2020 Aug 1. PMID: 32747076

11. Gururaj N, Voruganti N. The Prevalence of low T3 syndrome in chronic heart failure:A Hospital-based study. PERSPECTIVES IN MEDICAL RESEARCH [Internet]. Prathima Institute of Medical Sciences; 2021 Apr 6;8(3):20–4. Available from: http://dx.doi.org/10.47799/pimr.0803.05

